# Perceptual constancy of pareidolias across paper and digital testing formats in neurodegenerative diseases

**DOI:** 10.1101/2024.02.08.24302504

**Authors:** Gajanan S. Revankar, Tatsuhiko Ozono, Maki Suzuki, Hideki Kanemoto, Kota Furuya, Kazue Kamae, Kenji Yoshiyama, Yuki Yamamoto, Issei Ogasawara, Natsuki Yoshida, Susumu Iwasaki, Chizu Saeki, Yoshiyuki Nishio, Daisaku Nakatani, Kanako Asai, Yuta Kajiyama, Mikito Shimizu, Tatsuya Hayashi, Seira Taniguchi, Yu Suzuki, Rino Inada, Tomoya Taminato, Yoshitaka Nagai, Mamoru Hashimoto, Manabu Ikeda, Etsuro Mori, Hideki Mochizuki, Ken Nakata

**Author notes:** Correspondence: Gajanan S. Revankar, Center for Global Health, Dept. of Medical Innovation, Graduate School of Medicine, Osaka University Hospital 2-1, Yamadaoka, Suita, Osaka, Japan – 5650871.

## Abstract

Pareidolias refer to visual perceptual deficits where ambiguous shapes take on meaningful appearances. In neurodegenerative diseases, pareidolias are examined via a paper-based neuropsychological tool called the noise pareidolia test. In this study, we present initial findings regarding the utilization of pareidolia test on a digital format to analyze variations between paper-based and digital testing approaches. We performed our experiments on healthy controls, patients diagnosed with Alzheimer’s disease (AD), Dementia with Lewy body disease (DLB) and Parkinson’s disease (PD). Baseline MMSE assessments were conducted, followed by pareidolia testing using both paper-based tools and smartphones. Bland-Altman analysis was performed to evaluate the agreement between the two methods. We found that the illusionary phenomenon of pareidolia is consistent across paper and digital modalities of testing; that perceptual constancy is maintained across patient groups despite variations in image sizes; and pareidolic misperceptions, to some extent, are stabilized on a digital format. Our findings demonstrate a practical way of testing pareidolias on smartphones without compromising on the functionality of the test.

## Introduction

Pareidolias are visual misperceptions wherein ambiguous forms appear meaningful^1^. These perceptual errors are a natural phenomenon occurring in normal healthy individuals and reflect a transient mismatch between bottom-up sensory information and internally generated top-down visual processing^2^. Pareidolias can manifest in various forms, of which face-pareidolia stands out as the most prevalent and consistently defined phenomenon in literature^3^.

Although pareidolias appear innocuous among the healthy, they serve as important markers for neuropsychiatric disorders. Clinically, with the loss of insight, pareidolias act as early behavioral markers for psychosis in Alzheimer’s disease (AD)^4,5^, Dementia with Lewy body disease (DLB)^6,7^ and Parkinson’s disease (PD)^8–10^ establishing it as a significant prognostic indicator for disease progression^7^.

Pareidolias are quantitatively evaluated using the Noise pareidolia test (NPT), a simple, paper-based neuropsychological test comprising of black-and-white images of noise with faces embedded in them^11^. The NPT prompts a visuo-perceptual demand, necessitating the redirection of attention based on the sensory prominence displayed by the target stimuli^6,12^. We previously demonstrated neural correlates that affect frontal cortex network and attentional dynamics in PD patients using the NPT on a PC monitor screen, with screen dimensions like that of a paper-test^12,13^. Nevertheless, it remains uncertain whether alterations in the viewing distances or sizes of these images impact visual perception, especially in the absence of any eye-related pathologies.

It is known that our perception maintains the size of an object as relatively constant, even when there are alterations in the size, shape, or color of its retinal image due to variations in viewing distance^14,15^. This ‘perceptual constancy’ is suggested to be affected in neurodegenerative disorders like AD, PD, DLB or frontotemporal dementia, dependent much on the temporal course of the disease^10,16–18^. Since pareidolias affects visual processing encoded at multiple cortical levels at different stages of the disease^8^, we posited that perceptual tasks necessitating high-level vision and object recognition may affect perceptual constancy.

The aim of the study was, therefore, to clarify whether patients with neurodegenerative disease maintain a stable perception of NPT despite variations in image size and distance on different surfaces. To that end, we performed the NPT on two formats – one on paper and the other on a smartphone and outlined the qualitative and quantitative differences between the two. Practically, the outcomes of this study will allow smartphone-based evaluation of pareidolias in terms of clinical research.

## Methods

### General information

11 Healthy controls (HC), 30 AD, 26 DLB and 5 PD participants were prospectively enrolled for the study (Study start: Nov 2022). We recruited patients in their early to moderate stage of the disease. HC were sampled from a single center, and patients were recruited from 3 different hospitals across Osaka, Japan. Inclusion criteria for patients were (i) ≥ 40 years of age, and (ii) diagnosed as AD, DLB or PD according to their respective clinical diagnostic criteria^19–21^.

Patients with any of the following conditions were excluded: (i) if the attending physician/experimenter judged if a patient had severe behavioral or motor impairment hampering the usage of a smartphone, (ii) known eye-related pathologies, (ii) those on antipsychotic medications (olanzapine, risperidone, clozapine), (iii) major psychiatric diagnosis that affected activities of daily living, and (iv) uncontrolled major medical illness such as seizures or cardiovascular diseases. Healthy participants HC were ≥ 40 years of age, without any past or present neurological problems. Participants were tested with a near-vision Snellen chart integrated in the smartphone to include normal or corrected-to-normal vision subjects only. All participants provided written informed consent. The Osaka University institution review board cleared the protocol for the study to be performed in the Department of Neurology and the Department of Psychiatry, Osaka University, Nippon Life Hospital and Asakayama General Hospital all located in Osaka, Japan in accordance with the ethical standards of the Declaration of Helsinki (IRB Approval number – 22307). Clinical and neuropsychological assessments were conducted by a clinical psychologist or a neurologist, all performed in a single out-patient visit.

### Experiment flow

Following informed-consent, participants underwent 3 tests in the following order – i) Paper Noise pareidolia test (pNPT)^11^, ii) Mini-mental State Examination, Japanese version (MMSE-J)^22^ and iii) the smartphone Noise pareidolia test (sNPT). The pNPT comprises of 40 black and white images, with a face embedded in 8 of the 40 images. Participants must locate and identify faces accurately, and any misidentification of noisy areas as faces are marked as pareidolia. The images on pNPT were administered on A4-sized pages, placed flat on a desk with the participant viewing these images at approx. 40 to 50cms. Participants needed to exhibit 2 or more pareidolias in the test to be classified as pareidolia positive.

Pareidolia images in sNPT were the same as pNPT but were randomized to prevent a learning effect, although this effect is known to be minimal or non-existent^23^. Participants performed the sNPT test on an Android smartphone [Samsung A32 with screen dimensions 164mm (h) x 76mm (w), portrait mode], that was fixed on a smartphone mount, with participants sitting at 20-30 cm from the screen. Screen resolution and brightness were maintained at a constant level across all participants. Although participants were motivated to perform the sNPT independently, examiners aided when requested. The total test time for all the 3 tests were approx. 30 min.

### Data Collection and Statistical analysis

All clinical data were stored in paper-based case report forms. Digital data from sNPT were saved on a database in a cloud server (AWS) with the processing and readouts done offline. Statistical analysis was performed on JASP (version 0.18.2). Outcome variables included pareidolia scores (false-positives), missed responses (false-negatives) and correct responses (true-positives and true-negatives) for pNPT and sNPT. To report significant differences between groups, the statistical value was set to p < 0.05.

For 2-group comparison, Welch’s t-test or Mann Whitney-U test were performed depending on satisfactory assumptions of normality. For correlational analysis, Spearman’s rho coefficient was reported for continuous data, and Chi-squared tests for binary/categorical variables. Due to the nature of NPT, pareidolia scores are almost always positively skewed distributions^12,23^. Scores were therefore log-transformed for Bland-Altman (BA) analysis to evaluate the agreement between paper and digital methods^24^. A mean bias line for BA plots were shown to identify systematic difference between the measurement methods. Limits of Agreement (LOA) were set at 2 SD’s of the mean difference^25^ and confidence intervals reported for both mean bias line and LOA’s. A regression line (proportional bias) was plotted to ascertain whether the bias remained consistent across the test.

Data analyzed in this study will be made available from the corresponding author upon reasonable request. The paper version of NPT is available under an open license for research use. The software code used for this work has dependencies on internal tooling and infrastructure, is under patent protection (application number: JP2022-179766). The data are not publicly available due to privacy or ethical restrictions.

## Results

### General overview

72 participants (11 healthy controls (HC) and 61 patients; 34F and 38M) were consecutively enrolled. Their characteristics are summarized in Table-1. One AD and one DLB patient could not complete the digital version (sNPT) of the test due to fatigue and were excluded from analysis. One HC exceeded the cut-off for pareidolia score (=2).

**Table 1.**
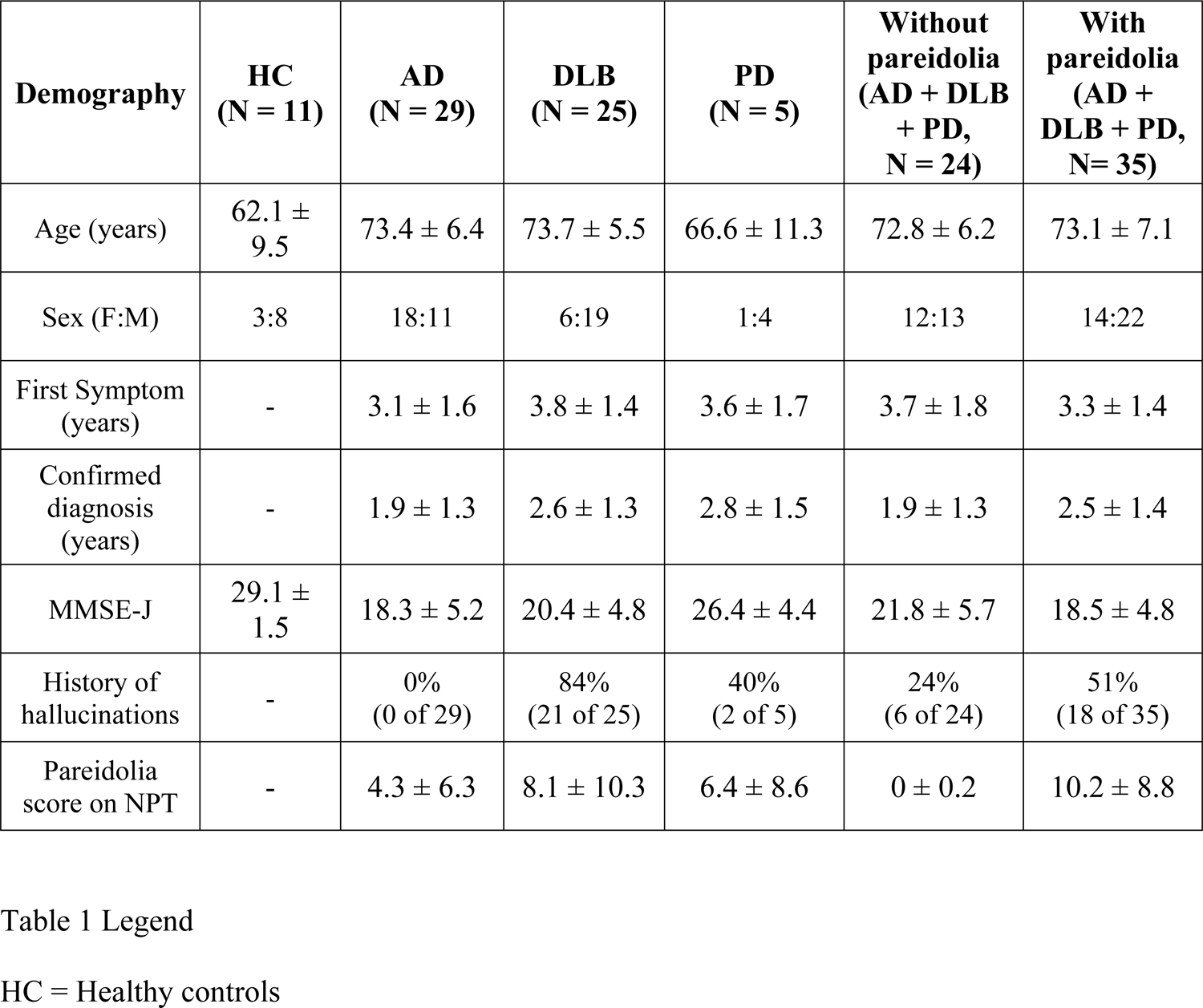
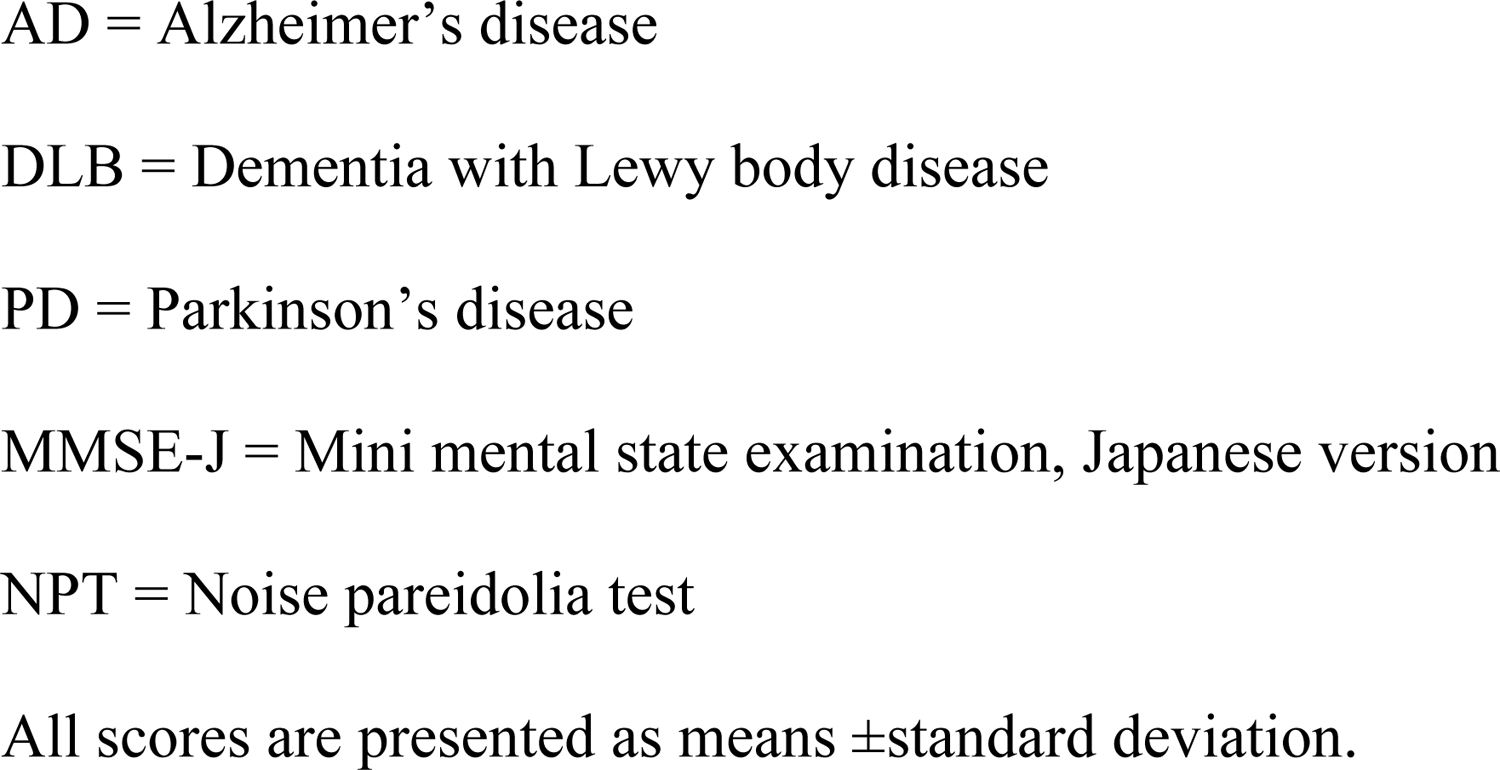
Participant characteristics.

| Demography | HC<br>(N = 11) | AD<br>(N = 29) | DLB<br>(N = 25) | PD<br>(N = 5) | Without<br>pareidolia<br>(AD + DLB<br>+ PD,<br>N = 24) | With<br>pareidolia<br>(AD +<br>DLB + PD,<br>N= 35) |
| --- | --- | --- | --- | --- | --- | --- |
| Age (years) | 62.1 ± 9.5 | 73.4 ± 6.4 | 73.7 ± 5.5 | 66.6 ± 11.3 | 72.8 ± 6.2 | 73.1 ± 7.1 |
| Sex (F:M) | 3:8 | 18:11 | 6:19 | 1:4 | 12:13 | 14:22 |
| First Symptom<br>(years) | - | 3.1 ± 1.6 | 3.8 ± 1.4 | 3.6 ± 1.7 | 3.7 ± 1.8 | 3.3 ± 1.4 |
| Confirmed<br>diagnosis<br>(years) | - | 1.9 ± 1.3 | 2.6 ± 1.3 | 2.8 ± 1.5 | 1.9 ± 1.3 | 2.5 ± 1.4 |
| MMSE-J | 29.1 ± 1.5 | 18.3 ± 5.2 | 20.4 ± 4.8 | 26.4 ± 4.4 | 21.8 ± 5.7 | 18.5 ± 4.8 |
| History of<br>hallucinations | - | 0%<br>(0 of 29) | 84%<br>(21 of 25) | 40%<br>(2 of 5) | 24%<br>(6 of 24) | 51%<br>(18 of 35) |
| Pareidolia<br>score on NPT | - | 4.3 ± 6.3 | 8.1 ± 10.3 | 6.4 ± 8.6 | 0 ± 0.2 | 10.2 ± 8.8 |

HC = Healthy controls
AD = Alzheimer's disease DLB = Dementia with Lewy body disease PD = Parkinson's disease MMSE-J = Mini mental state examination, Japanese version NPT = Noise pareidolia test All scores are presented as means ± standard deviation.

Across patient groups [N=59, Aged= 73yr ±6.7, (Mean, SD)], at the time of our testing, the duration of first appearance of symptoms was 3.5yrs ±1.5 and the duration of getting a definitive clinical diagnosis using the diagnostic criteria was 2.3yrs ±1.4. Approx. 60% (35 of 59) of the evaluated patients exhibited pareidolias on the pNPT, and around 51% of them had a history of hallucination. The presence of pareidolia on pNPT showed a positive correlation to history of hallucinations (*X*^2^ (1, N=59) = 4.09, *p = 0.043*).

Pareidolia scores correlated negatively with MMSE (r= −0.37, *p=0.004*), but not to age (r= 0.01, p=0.93) or disease duration / first symptom (r= 0.03, p=0.80). Patients with pareidolias had a 3-point lower MMSE score compared to those without pareidolia (U=584.0, *p=0.011*, rank biserial effect size = 0.39). In DLB patients, the pareidolia score exhibited significant correlations with MMSE scores in comparison to AD (Supplementary figure - 1).

### Paper vs Digital testing

Figure-1A provides a summary of correlational measures for test outcomes between paper (pNPT) and digital measurements (sNPT) for 70 participants (HC=11 and patients=59). Scores correlated significantly for pareidolias, missed images and correct answers (true positive + true negatives) variables.

**Fig 1.**
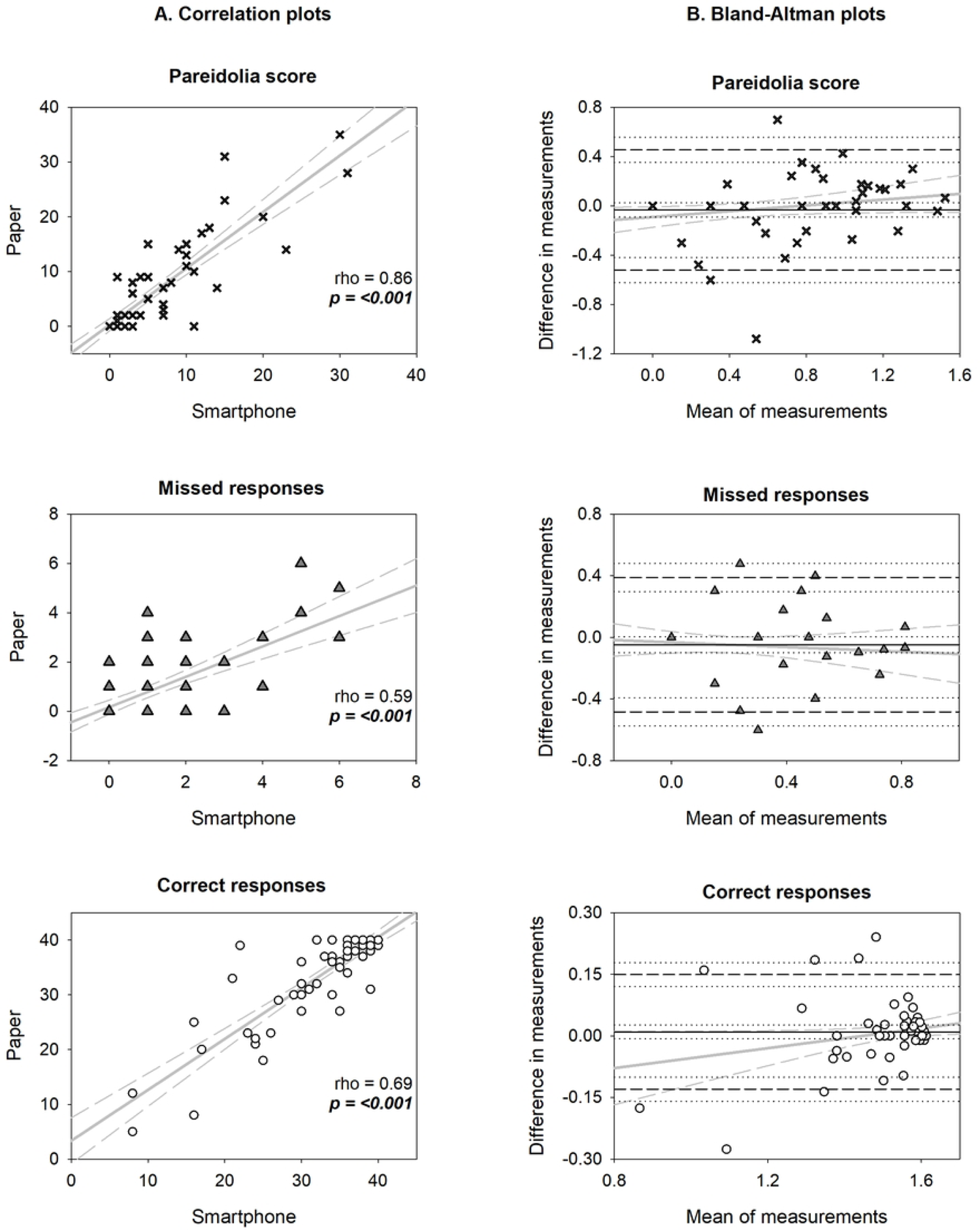
Outcome measures of pareidolia test performed on paper versus smartphone. A. Shows a column of correlation plots for pareidolia, missed and correct responses between the 2 modalities. Regression line is shown in solid gray, with confidence intervals (CI) in dashed gray. Spearman’s rho and p values are defined within the graphs. B. Bland Altman plots column show the mean and difference of measurements in x and y axes respectively following log transformation of raw data. Solid black line represents the mean bias with dashed black lines representing the limits of agreement (LOA) (2SD’s). Black dotted depict CI’s for the bias and LOA. Solid gray and dashed gray lines illustrate the proportionality bias.

BA plots (Figure-1B) indicated strong agreement between pNPT and sNPT, with the confidence intervals (CI) falling within the mean bias line close to zero for all the 3 outcome variables. Furthermore, over 95% of the values for all three variables were within the limits of agreement (LOA) (Table-2).

**Table 2.** BA values for outcome variables.

| Responses | Bias<br>(CI's) | Lower<br>LOA | Upper<br>LOA | BA statistic | p-value |
| --- | --- | --- | --- | --- | --- |
| Pareidolia | -0.03<br>(-0.09 to 0.02) | -0.52 | 0.45 | t = -1.1, dF =69 | 0.30 |
| Missed | -0.05<br>(-0.10 to 0.00) | -0.48 | 0.39 | t = -1.8, dF =69 | 0.08 |
| Correct | 0.01<br>(-0.00 to 0.03) | -0.13 | 0.15 | t = 1.1, dF =69 | 0.30 |

For pareidolias, a positive trend was observed proportional to the magnitude of their scores, i.e. a greater spread in the measurements between pNPT and sNPT for low and mid-range scores. High pareidolia scores remained unaffected between the measurement modalities. Variability was also consistent across the graph with the scatter around the bias line being fairly constant. Missed responses generally did not show major trends and were similar between pNPT and sNPT. For lower levels of correct responses, the agreement limits seem to widen, while the bias decreases significantly for higher levels of correct responses, characterized by a dense scatter.

Intuitively, following BA analysis, we binned pareidolia-positive patients into 3 distributions, ≥2 and ≤4, ≥5 and ≤10, and ≥11 pareidolias with 11, 12 and 11 samples respectively. A visual representation of this effect is shown in Figure-2 which demonstrates a tighter distribution for sNPT compared to pNPT. Levene’s test showed a significant difference of variances between sNPT and pNPT groups for low and mid-range pareidolia scores.

Fig 2. shows raincloud plots for pNPT and sNPT represented in green and orange respectively. The differences between pNPT and sNPT scores across 3 distribution bins are described via means and SD’S shown above the box plots. For ≤10 pareidolias on sNPT, the variances were much less compared to pNPT. However, ANOVA tests did not achieve statistical significance for the binned groups.

**Fig 2.**
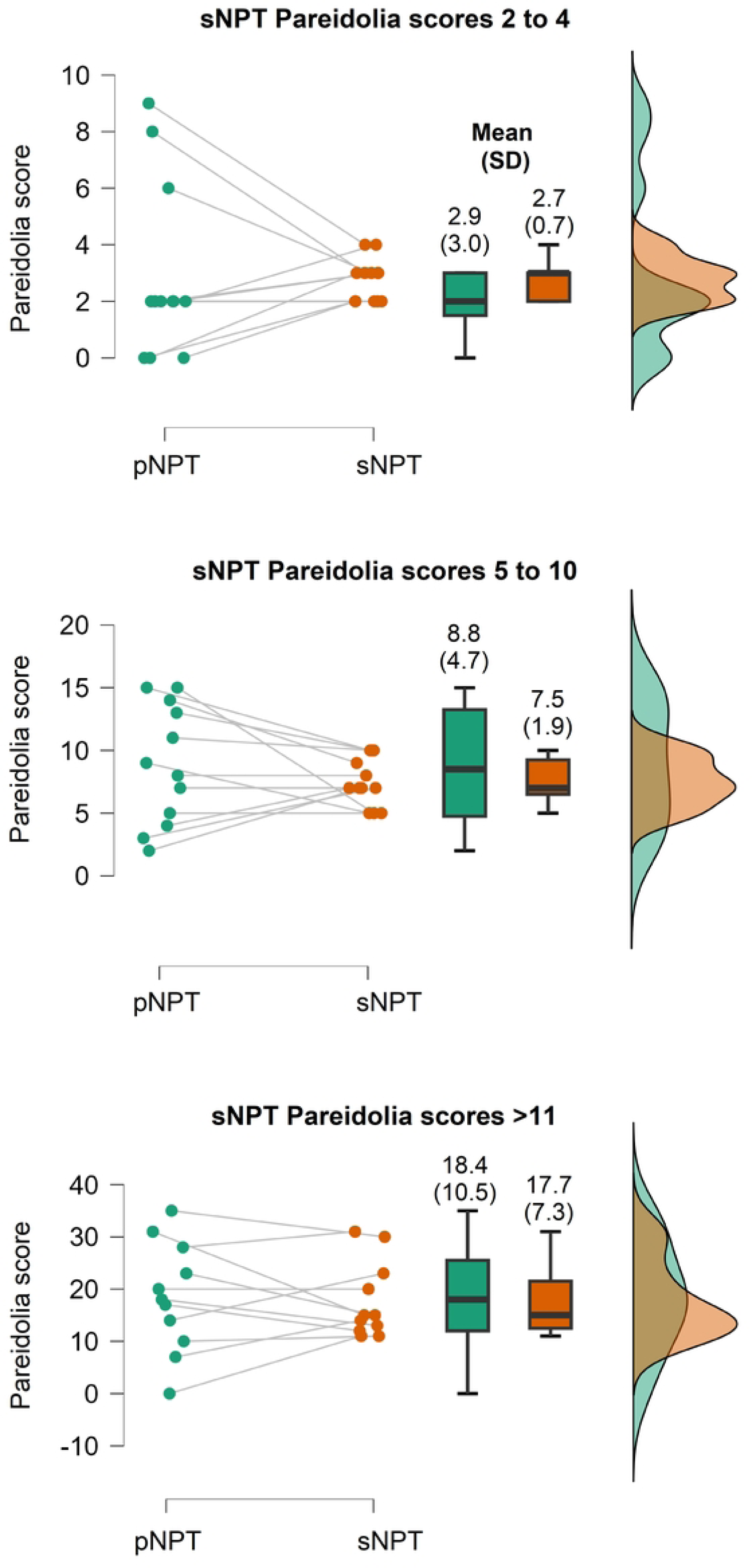
Distribution of pareidolia scores between paper (pNPT) and smartphone (sNPT)

## Discussion

We investigated whether pareidolic perception is altered in patients with neurodegenerative diseases when tested across different-sized formats. We found that: (i) the visuo-perceptual phenomenon of pareidolia is consistent across paper and digital modalities of testing, (ii) size constancy is maintained across participant groups despite variations in image sizes, and (iii) pareidolic misperceptions, to some extent, are stabilized on a digital format.

Prior studies have reported pareidolias to occur in up to 30% of individuals with AD^4,5^, around 50% of those with PD^8,9^, and up to 80% of individuals diagnosed with DLB^6^, establishing it as a significant prognostic indicator for disease progression^7^. We noticed similar characteristics among our group of patients, along with a considerable delay in confirming a definitive diagnosis (around 14 months), and a connection between hallucinations and worsening cognitive abilities in the presence of pareidolias.

In this study, our primary goal was to verify the potential utilization of digital pareidolia (sNPT) testing and address any differences with paper-based methods (pNPT). Pareidolia quantification on the NPT is an active search-and-detect paradigm requiring integration of both bottom-up sensory input with a stronger top-down, knowledge-based modulation^1,12^. Illusionary responses to the NPT may further manifest in several forms - of objects, of animals and so on - although a greater attentional demand is required for preferential selection of faces. Furthermore, pareidolias are highly context dependent and represent an endogenous bias in patients^6,7^. With respect to these mechanistic characteristics, we speculate that the brain employs a combination of perceptual and cognitive mechanisms, including top-down processing, contextual cues, and size constancy, to interpret and maintain consistency across different surfaces despite variations in image size or distance. This “stable misperception”, without any visual pathologies, might result from factors other than perception itself. Such factors could include dysfunctional attentional control^12^, a lack of insight with abnormal inferences^6^, and suggestibility^7^, characteristic of AD and DLB pathophysiology^26^ which could be device agnostic (e.g. smartphones, monitors etc). Our former studies incorporating NPT on a 20” monitor screen^12,13^ and the current experiment on 3”x3” smartphone screens strengthens this assertion.

Another observation we made in the digital format was that the scores for pareidolia on the sNPT showed a reduced dispersion compared to the pNPT, particularly noticeable for both low and mid-range pareidolia scores in the sNPT. This effect could be attributed to several factors. Performance differences have been reported in studies reviewing reading and comprehension paradigms between display monitors and on paper, although the effects are inconclusive, and one may not be better than the other^27,28^. Reallocation of visual processing resources from perceptual to cognitive domains (working memory, executive function) is suggested to affect performance in such modalities^28^. We speculate device-related changes in contrast sensitivity, resolution and brightness may affect sensory inputs impacting visual processing pathways^29,30^. Secondly, with respect to the study design, sNPT was always performed after pNPT. It is likely that some form of adaptation in terms of individual preferences (use of smartphone) and prior experience may have affected user performance^23^.

### Limitations

Isolation of early levels of visual processing such as color vision, stereoacuity, contrast sensitivity may not have been adequately characterized in this study. The severity of dementia among patients were also not strictly controlled (MCI-level, moderate-level, and so on) as the patients were consecutively enrolled. At the time of testing, most patients were on Donepezil and/or Levodopa with a combination of other non-neurological medication. The effect of medication on visuo-perception should be carefully studied in the future. As observed in our sample, quantification of pareidolias alone do not have sufficient sensitivity to distinguish between different types of neurodegenerative diseases^5^. Given the current findings, it may be pertinent to reevaluate pareidolia cut-off scores that is specific for digital devices. We specifically evaluated a solitary visuo-perceptual phenomenon and correlation with other functional neuropsychological testing domains would be relevant. Within group differences (e.g. AD with and without pareidolias, etc.) may be of significant interest but were not formally evaluated because of low sample sizes. It would be justified to explore these aspects in future studies.

In conclusion, qualitative and quantitative differences of the NPT on different formats are minimal. The results of this study may open a practical / simpler way of testing pareidolias on smartphone format without compromising on the functionality of the test.

## Data Availability

Data cannot be shared publicly because of ethical restrictions. Data are available from the Osaka University Institutional Data Access / Ethics Committee (via email) for researchers who meet the criteria for access to confidential data.

## Acknowledgements

Our sincere thanks to the patients who participated in the study, to the Codemonk team for the software development and maintenance of the app. Our thanks to Dr. Ryo Takahashi, Dr. Manabu Sakaguchi, Dr. Junya Kobayashi, Dr. Seema Nachankar, Ms. Arya Revankar, Dr. Karen Fortuna, Dr. Vinamrita Singh, Ms. Julia Hill, Ms. Atsuko Masuda, and Mr. Nityanand Vernekar for their assistance with development in an earlier phase of the project.

## Supporting information

**S1 Fig.** Correlation between pareidolia and MMSE scores for AD and DLB

**S1 Fig.** shows data with regression line in solid black with CI’s in solid gray, with Spearman’s correlation coefficient values within the graph. MMSE-J = Mini-mental state examination, Japanese version. DLB patients had a strong negative correlation between Pareidolia and MMSE scores contributing significantly to the patient group.

## Author contributions

Conceptualization: GSR, MI, EM, HM

Methodology: GSR, TO, MS, HK, KF, YK, MH, MI, EM, HM, KN

Software: GSR, IO, CS, YK

Investigation: GSR, TO, MS, HK, KF, KK, KY, YY, IO, SI, CS, YN, DK, KA, NY, YK, MiS, TH, TT, ST, YS, RI, YNa, MH, MI, EM.

Validation: GSR, TO, YK, YNa, MH, MI, EM, HM, KN

Formal analysis: GSR, EM

Resources: GSR, TO, MS, HK, KF, KK, KY, IO, YN, DK, YN, MH, MI, EM, HM, KN

Data Curation: GSR, TO Writing - Original Draft: GSR

Writing - Review & Editing: TO, MS, HK, KF, KK, KY, YY, IO, SI, CS, YN, DK, KA, NY, YK, MS, ST, YS, RI, YN, MH, MI, EM, HM, KN

Visualization: GSR, EM Supervision: GSR, MI, EM, HM, KN Project administration: GSR

Funding acquisition: GSR

